# Deep Learning-Based Prediction Model of Heart Failure with Improved Ejection Fraction

**DOI:** 10.1101/2023.10.30.23297807

**Authors:** Yan Zhang, Yanbing Pan, Yanzi Zhang, Yongle Wang, Qiuting Jia, Yihui Kong

## Abstract

**Background:** This study developed a deep learning model to predict improvement of left ventricular ejection fraction in patients with heart failure.

**Methods:** An internal database comprising clinical, laboratory and echocardiographic features obtained from the First Affiliated Hospital of Harbin Medical University was used to construct and train the deep learning model.

**Results:** A total of 422 cases were included in this study. 122 (28.9%) were patients with HFimpEF, 300 (71.0%) were patients with HFrEF. Multivariable analyses showed that smaller baseline left atrial anterior-posterior diameter (LAD) and left ventricular end-diastolic diameter (LVEDD), higher baseline interventricular septal thickness at diastole (IVSD) and levels of prealbumin were the independent clinical predictors of LVEF improvement. Deep learning model demonstrated an overall predict accuracy of 96% in the validation set and 89% in the training set.

**Conclusions:** Independent predictors of LVEF improvement were smaller baseline LVEDD, LAD, higher baseline IVSD and baseline levels of prealbumin. Our deep learning model had shown acceptable performance in predicting improvement of left ventricular ejection fraction in patients with heart failure.

**Registration:** URL: https://www.clinicaltrials.gov; Unique identifier: NCT06070506.

## Introduction

Heart failure (HF) is a complex clinical syndrome in which various structural and/or functional abnormalities of the heart result elevated ventricular pressure and decreased cardiac output with corresponding signs and symptoms^1^. With high prevalence, rapid progression and poor prognosis, HF is a highly lethal and incurable clinical disease and a serious global public health problem.

Ventricular remodeling plays a key role throughout the course of HF, and the more severe the ventricular remodeling, the worse the prognosis for cardiovascular disease. Previous studies have found that, after pharmacological and device therapy, the process of adverse ventricular remodeling caused by activation of the renin-angiotensin-aldosterone and adrenergic nervous systems in patients with HF is reversible. The symptom manifests reductions in end-diastolic volumes, increased left ventricular ejection fraction (LVEF) and normalization of left ventricular volume and shape associated with improvement in both systolic and diastolic function. In particular, the improvement of LVEF can be up to 10% or more, from which the concept of heart failure with improved ejection fraction (HFimpEF) came, with differences in pathophysiological manifestations, clinical characteristics, and prognosis from other HF categories.

According to the 2022 American College of Cardiology and the American Heart Association and the Heart Failure Society of America (ACC/AHA/HFSA)^2^, HF can be classified into four categories:

- heart failure with reduced ejection fraction (HFrEF) with an EF ≤40%,
- heart failure with preserved ejection fraction (HFpEF) with an EF ≥50%,
- heart failure with mildly reduced ejection fraction (HFmrEF) with an EF between 41 and 49%,
- and heart failure with improved ejection fraction (HFimpEF) with previous EF ≤40% and a follow-up EF of more than 40%.

In recent years, deep learning algorithms have been widely implemented in the medical field to assist in diagnosis. Deep learning-based clinical profile can accelerate the diagnosis of HFrEF, HFpEF and HFmrEF patients^3^. However, the challenging remains in the diagnosis of HFimpEF requires multiple echocardiograms or cardiac magnetic resonance and laboratory data. Previous assessment addresses the differences in clinical characteristics between participants with HFrEF, HFpEF and HFmrEF. Until recently, there is some lack of methods about fast prediction of HFimpEF. The aim of this study was to design a deep learning-based trained model to assist in HFimpEF diagnosis.

## Methods

### Study population

The participants in the study are HF patients hospitalized in the Department of Cardiology of the First Affiliated Hospital of Harbin Medical University who had no less than two echocardiograms at baseline and during the follow-up period, between January 2014 to December 2022. Baseline data of the first admission from HF patients were obtained from Electronic Health Record system of the First Affiliated Hospital of Harbin Medical University.

Enrollment criteria is 18 years of age or older, and the diagnostic criteria of HF follows the 2018 Chinese Guidelines for the Diagnosis and Treatment of Heart Failure^4^, having symptoms of dyspnea, fatigue or decreased activity tolerance, having signs of fluid retention (such as pulmonary congestion and peripheral edema), having echocardiogram abnormalities in cardiac structure and/or function, showing elevated natriuretic peptide levels (BNP>35 ng/L or/and N-terminal pro-BNP >125 ng/L), reviewing echocardiography after discharge. Patients with hypertrophic, restrictive, or invasive cardiomyopathy and congenital or rheumatic heart disease, patients who had heart transplantation during follow-up were excluded.

Echocardiograms reviewed within 3∼12 months after discharge were collected, features including LVEF, LAD and left ventricular end-diastolic diameter (LVEDD). Based on the initial records and the check LVEF results, the patients have been divided into 2 groups: LVEF persistently ≤40% (HFrEF) and LVEF recovered to >40% (HFimpEF).

### Data selection

This study was approved by the First Affiliated Hospital of Harbin Medical University Medical Ethics Committee (2023JS19). Baseline data included demographic, clinical, laboratory data and echocardiographic features. Demographic and clinical information included age, sex, BMI, heart rate, pulse, New York Heart Association class (NYHA), HF etiologies, physical examination, comorbidities, smoking history, operation history, and etc. Laboratory data included erythrocyte, leukocyte, hemoglobin, platelet count (PLT), alanine aminotransferase (ALT), aspartate transaminase (AST), prealbumin, triglyceride (TG), total cholesterol (TC), low-density lipoprotein cholesterol (LDL-C), very low-density lipoprotein cholesterol (VLDL-C), high-density lipoprotein cholesterol (HDL-C), lipoprotein(a)[Lp(a)], fasting blood glucose, creatinine, uric acid, blood potassium, creatine kinase MB (CKMB), high-sensitivity cardiac troponin (hs-cTnI), BNP, NT-proBNP. Echocardiography measured by biplane Simpson method was used to assess LVEF, LVEDD, left atrial anterior-posterior diameter (LAD), interventricular septal thickness at diastole (IVSD).

## Deep learning model development

### Data preprocessing

Our deep learning model was trained on the NVIDIA GeForce GTX 1080 graphics processing unit (GPU) with 8 GB display memory (VRAM). Data preprocessing is used to improve the quality of raw data, facilitate deep learning model training, and promote the accuracy of the model. Before preprocessing, data need to be checked, and missing values will be refilled and normalized. The process is listed as below:

- Detect the missing value of data,
- remove the variables with missing data exceeding 1/3 of the sample size,
- fill in the missing data of the remaining variables with the median interpolation method,
- and normalize the 1D data to eliminate dimensional inconsistency.

### Deep learning model

Two-dimensional (2D) convolutional neural networks (CNNs) are extensively used to classify medical signals. Convolutional and pooling layers are used to concentrate 1D linear signals to discover certain disease characteristics. The two Conv2D layers are first introduced into the model. Through Conv2D layer, the original feature vector of HFimpEF is convoluted to form reasonable features which can be learned by other classifiers. Then the Batch Normalization layer is used to prevent the overfitting. ReLU activation function transfers the linear mapping into non-linear spaces. After that, max pooling layer is used to reduce the dimension of feature maps. Finally, the full connection layer consolidates all features and activate the classification with Softmax function to correctly classified data. The dropout layer was designed to randomly drop some connections between layers to prevent overfitting, and the dense layer was designed for output. As is shown in Figure 1, the network structure consists of 2 convolutional layers, 1 max pooling layer, 1 dropout layer and1 full connection layer.

**Figure 1.**
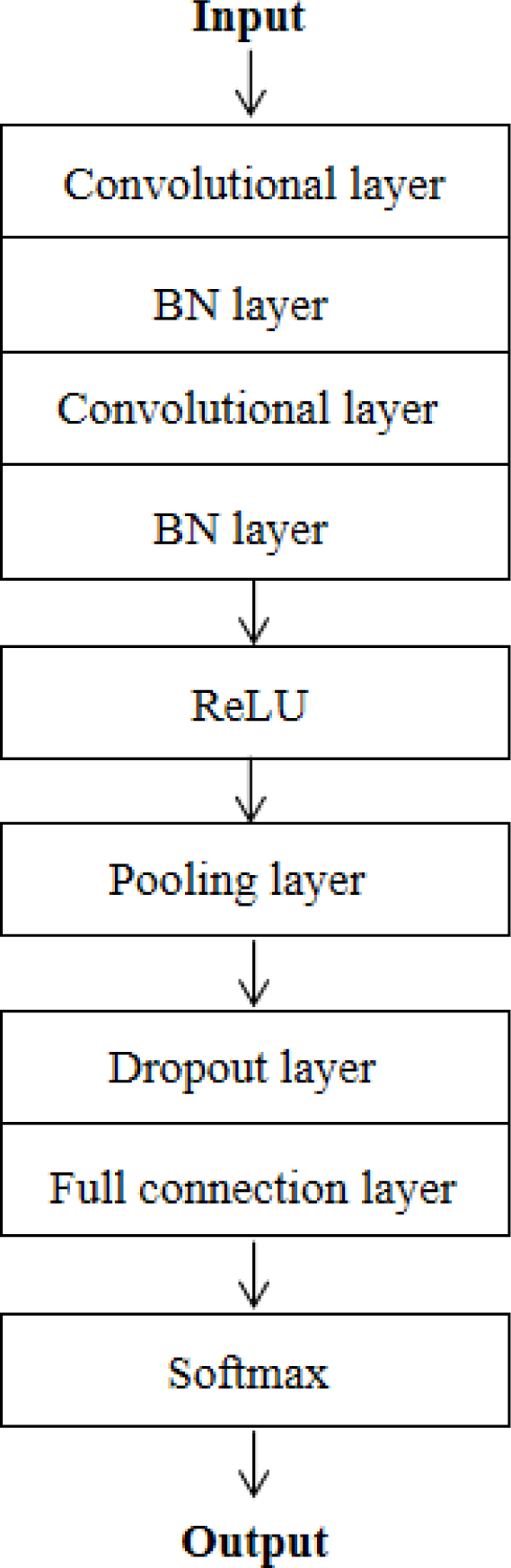
The Structure of Convolutional neural networks. The two convolutional layers are first introduced, then the Batch Normalization (BN) layer is used to prevent the overfitting. ReLU activation function transfers the linear mapping into non-linear spaces. After that, max pooling layer is used to reduce the dimension of feature maps, and the full connection layer consolidates all features and activate the classification with Softmax function to correctly classified data. The dropout layer was designed to randomly drop some connections between layers to prevent overfitting.

### Model training setup

The deep learning model should be trained before prediction. The original data are randomly distributed into training and validation data sets. From the training data set, 10% were randomly selected during the training process for 10-fold validation. The validation set is used to adjust the hyper-parameter and evaluate the generalization performance of the trained model. Adam is used as the optimizer. The initial learning rate is set to 1e-2. The mini-batch size is set to 32. The max epoch is set to 50 with early stopping and the dropout ratio is set to 0.2. The models take a series of samples as its inputs and give its classification result as output.

### Statistical analysis

SPSS version 25.0 has been used in statistical analysis. Values were presented as mean and standard deviation for continuous variables, and as numbers and percentages for categorical variables. Demographic, clinical, laboratory and echocardiographic features were compared between two groups with standard statistical methods including independent samples *t*-test for continuous variables and chi-square or Fischer’s exact test for categorical variables.

Logistic regression model was used to analyze the predictive factors of LVEF improvement. *P* value of <0.05 was considered statistically significant. For evaluation of the 2D CNN model, confusion matrix, accuracy, precision, recall and F1 score commonly applied in the evaluation of deep-learning models, were used.

## Results

### Study participants

A total of 1855 HF patients with echocardiogram were comprised in the study. After selection and exclusion, the internal data set finally included 422 patients containing 300 patients (70.1%) with HFrEF and 122 patients (28.9%) with HFimpEF (Figure 2).

**Figure 2.**
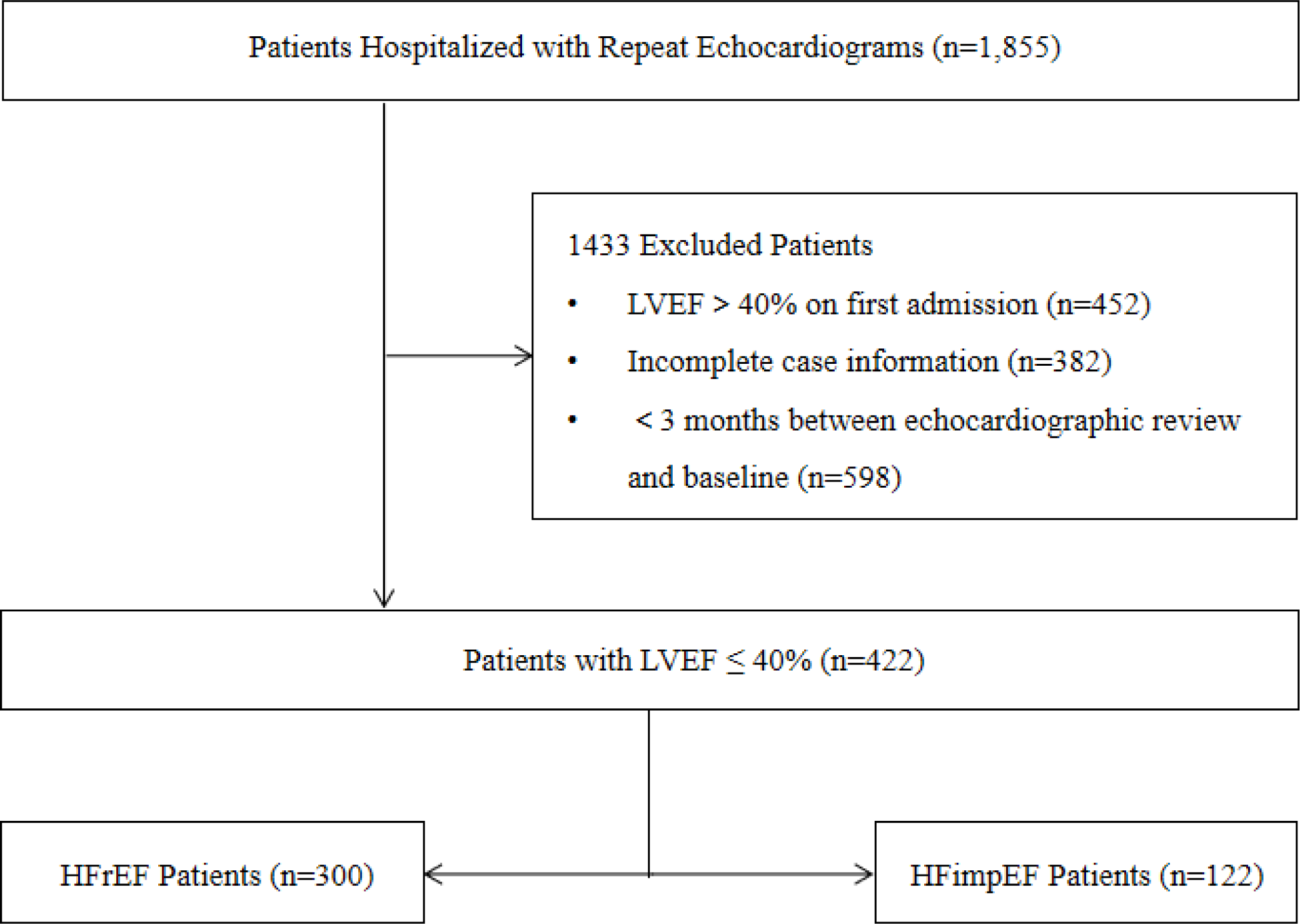
The Selection Flow Chart. A total of 1855 HF patients with echocardiogram were comprised in the study. After selection and exclusion, the internal data set finally included 422 patients containing 300 patients (70.1%) with HFrEF and 122 patients (28.9%) with HFimpEF.

### Baseline Characteristics

The baseline characteristics are presented in detail in Table 2. Overall, average age is (65.3±13.9) years and 65.2% patients are males. Average body mass index is (25.3±4.8) kg/m^2^. Briefly, hypertension was reported in 42.4% of patients, diabetes mellitus in 29.3%, dilated cardiomyopathy in 19.8% and coronary heart disease in 67.4% of patients of the study population. Average LVEF was (31.7±6.3) %, average LAD was (44.7±5.8) mm, median LVEDD was 43mm and median IVSD was 9.4mm.

**Table 1.**
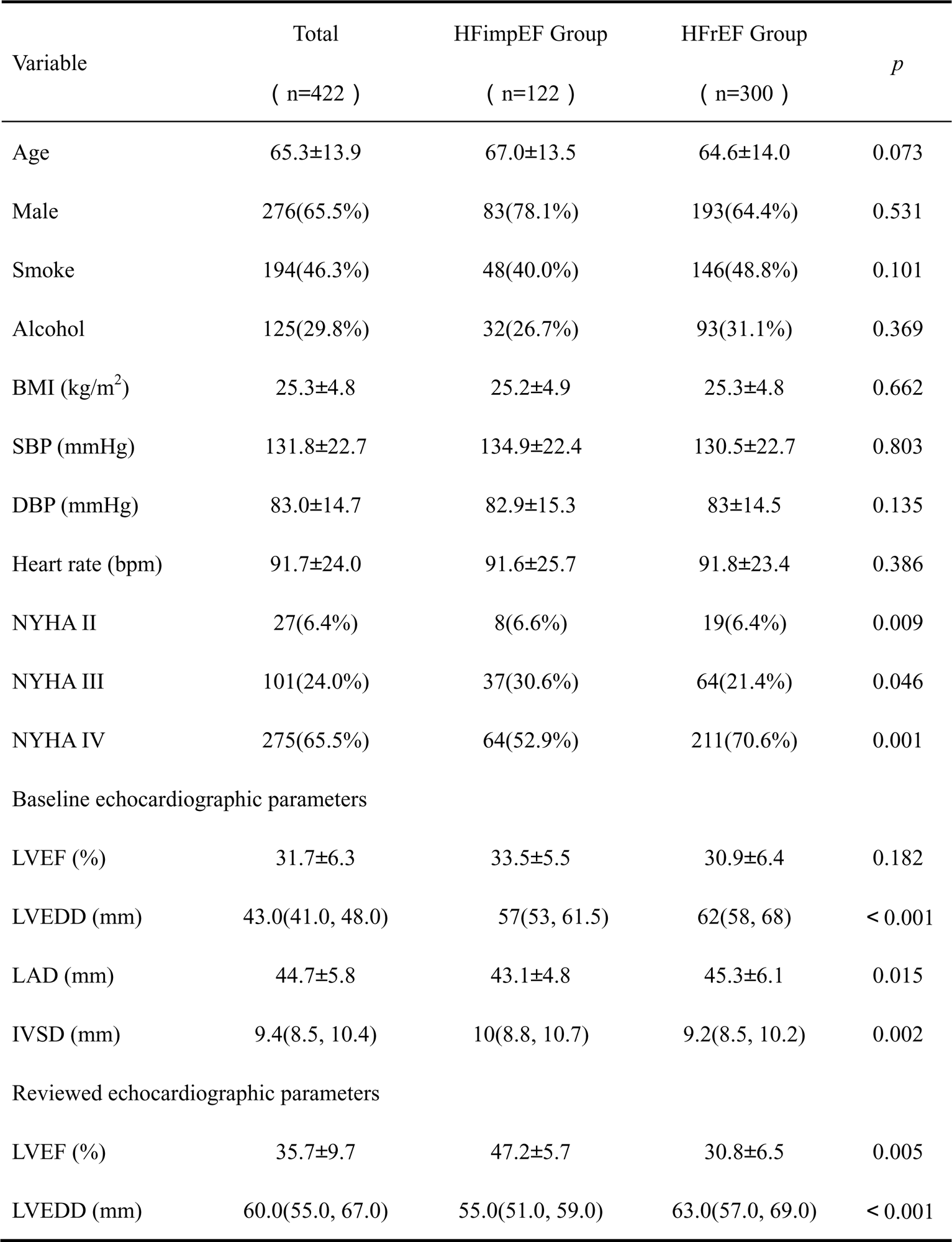
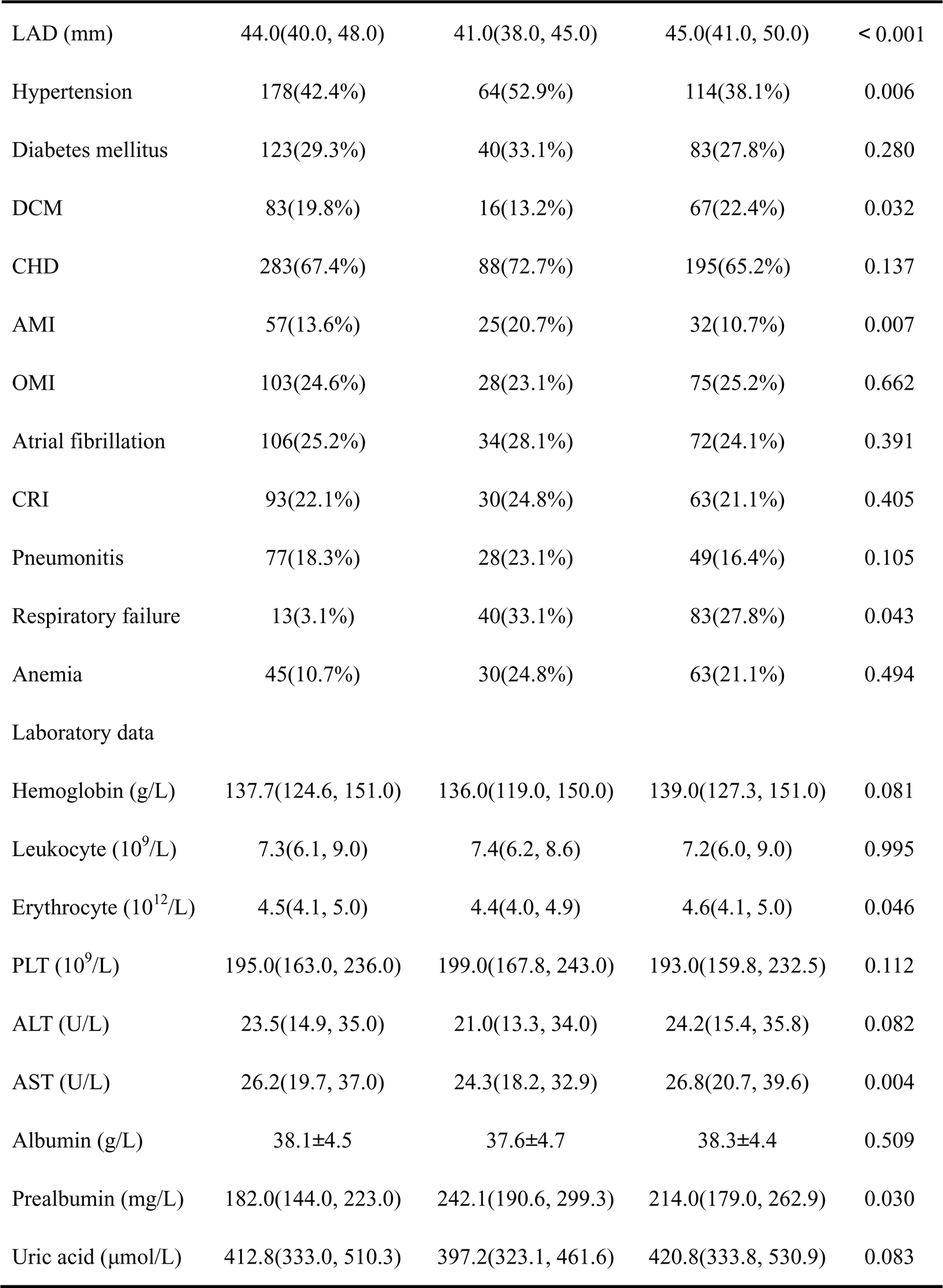
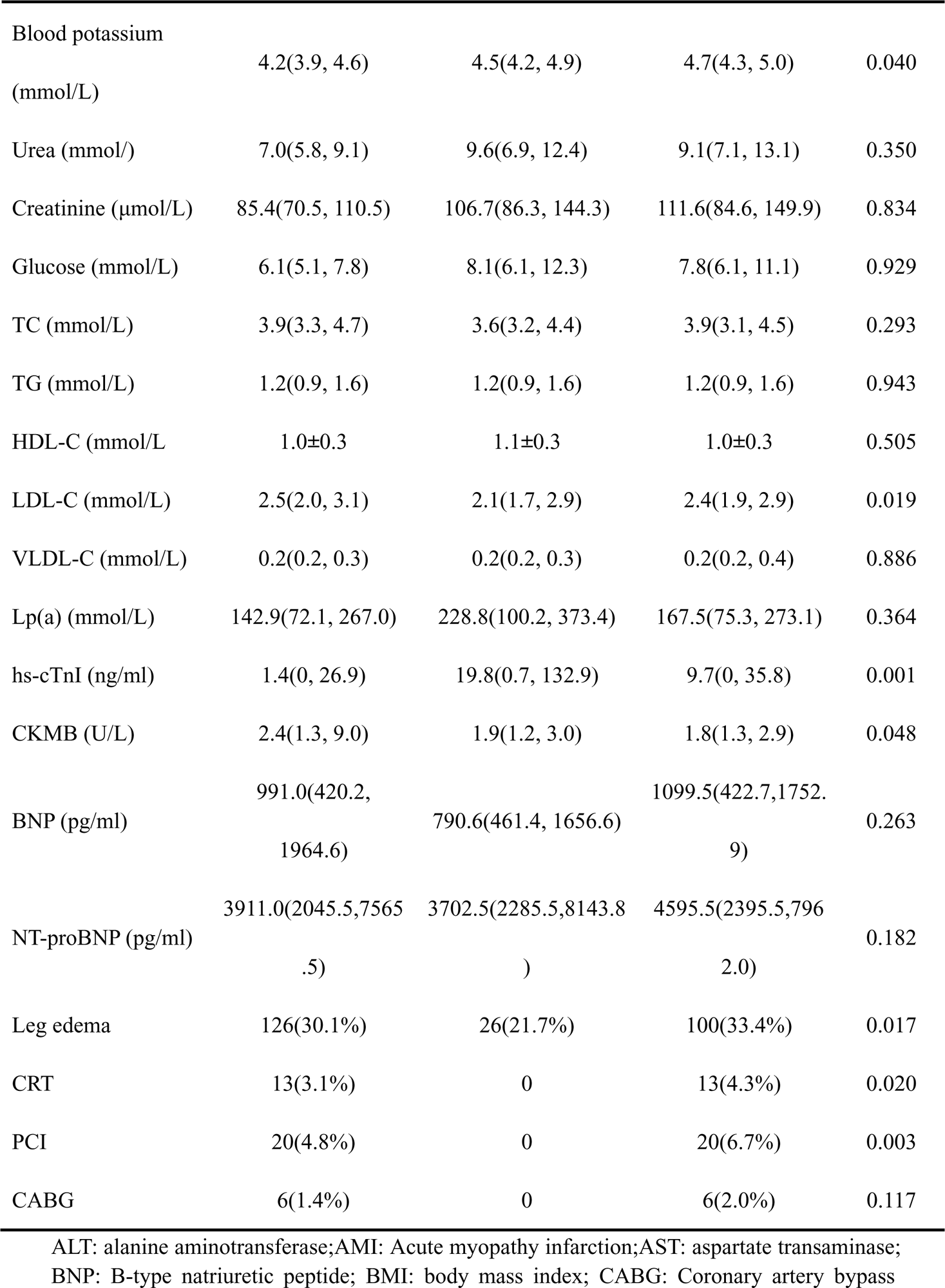
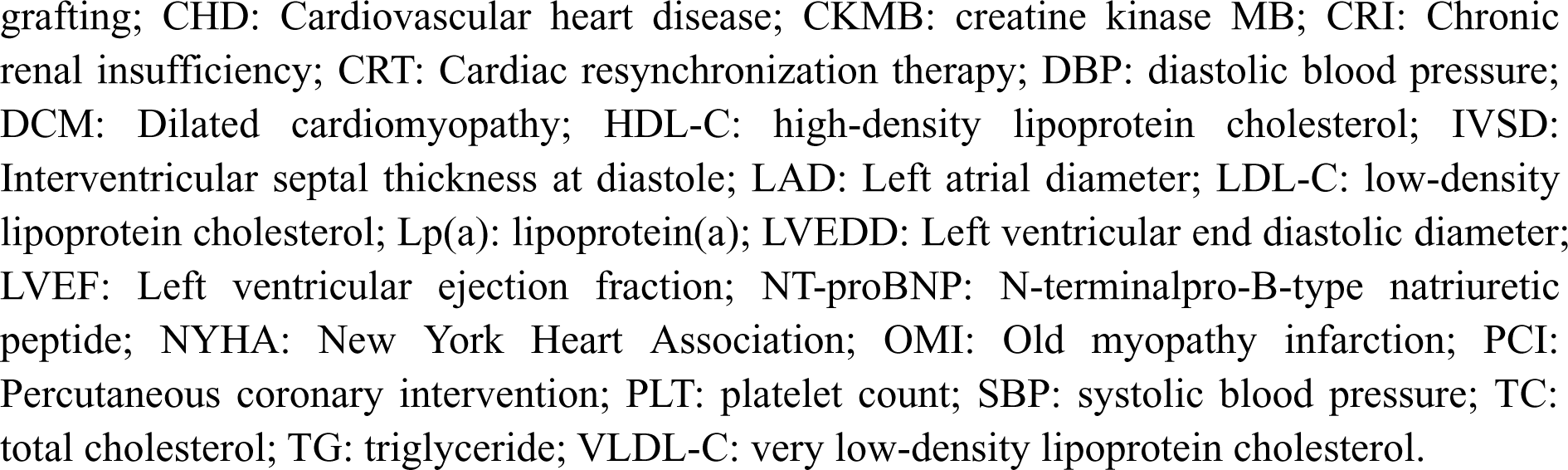
Baseline characteristics.

**Table 2.**
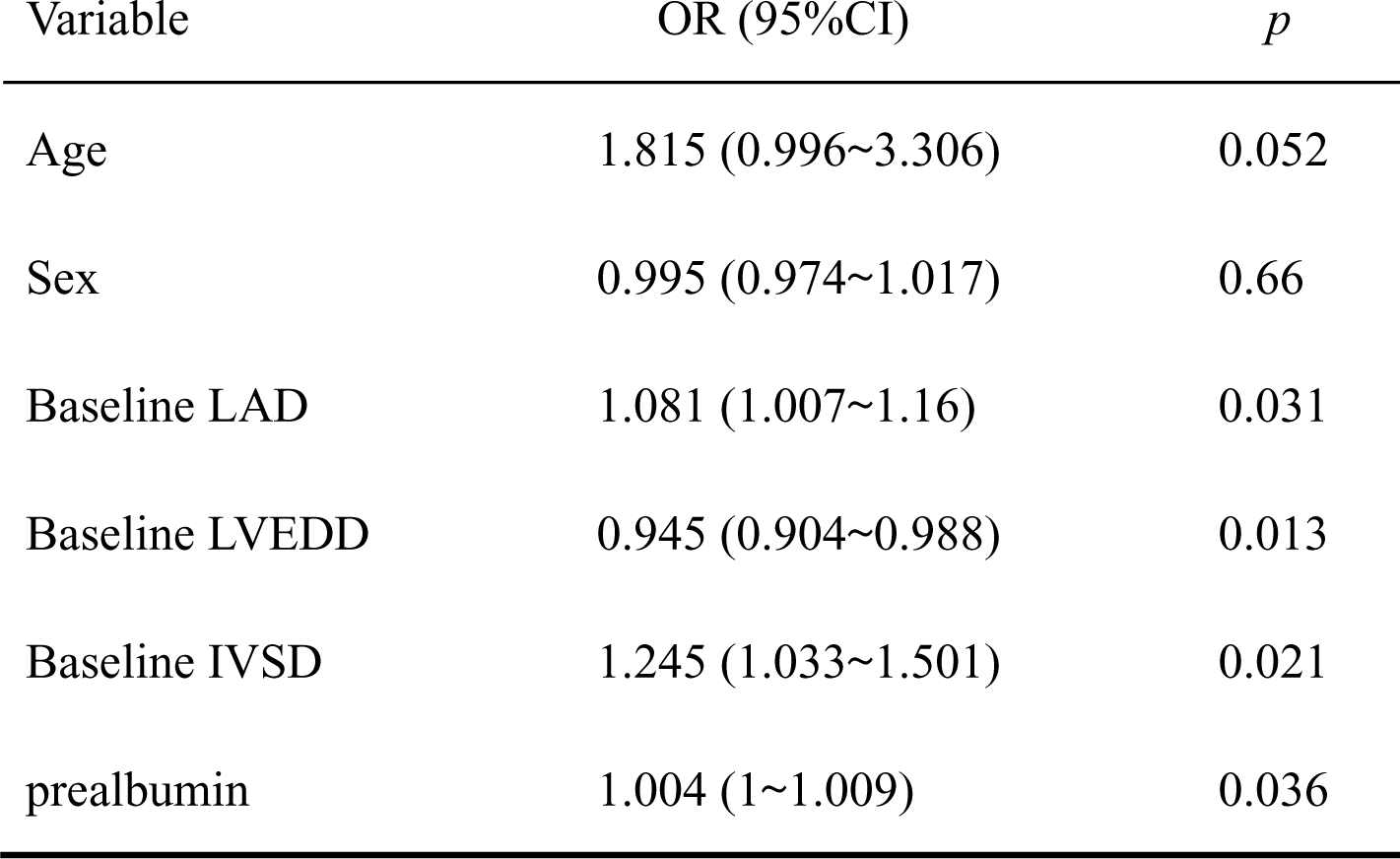
Multivariate analysis of variables associated with LVEF improvement.

Comparing these baseline characteristics, the most significant differences are that patients with HFimpEF have more patients with hypertension and acute myocardial infarction, less patients with dilated cardiomyopathy and combined lower limb edema, better functional class, higher hs-cTnI, CKMB and prealbumin levels and lower erythrocytes, potassium, LDL-C, and AST levels than HFrEF. Comparison of echocardiograms shows that patients with HFimpEF have lower baseline LAD and LVEDD values, higher IVSD values than HFrEF.

There is no significant difference in baseline LVEF between the two groups. Compared with baseline echocardiograms, patients with HFimpEF have significantly higher LVEF. The mean LVEF of HFimpEF increased from (33.5±5.5) % to (47.2±5.7) %. LAD and LVEDD values on review echocardiograms of patients HFimpEF decreased. On the contrary, an increasing in LVEDD is observed in the HFrEF group.

### Predictors of LVEF Improvement

In the univariate analyses, hypertension, acute myocardial infarction, dilated cardiomyopathy, NYHA class III/IV, baseline LVEF, LAD, LVEDD and IVSD, hemoglobin, erythrocyte, prealbumin, uric acid, potassium, and LDL-C were associated with the improvement of LVEF. In the multivariable analyses after adjustment of gender and age, smaller baseline LAD and LVEDD, higher baseline IVSD and prealbumin levels were the independent clinical predictors of LVEF improvement (Table 2).

### Deep Learning Predictive Modeling

Of 422 patients, 380 patient data have been finally randomized into a training data set and an internal validation data set with 42 patients. The whole model training takes 7s. Our model has a high precision level in HFimpEF prediction with an 89% in the training set and a 96% in the validation set (Figure 3). Validation accuracy is obviously higher than training accuracy, indicating that our model does not occur any adverse fitting. Furthermore, our model achieved high performance in accuracy, sensitivity, specificity, precision, and F1 score (Table 3).

**Figure 3.**
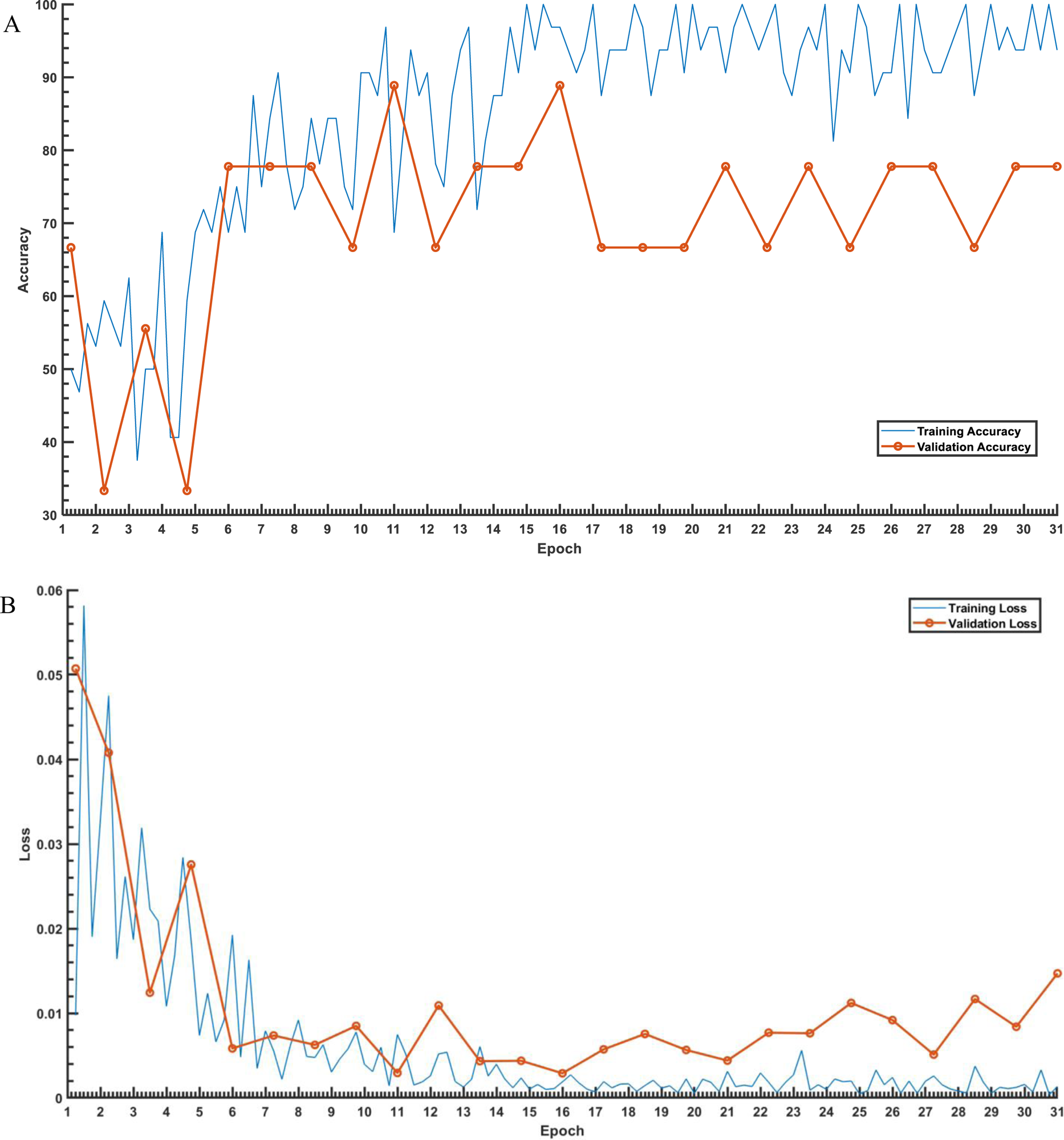
Accuracy-loss curve of training process. **(A)** Accuracy curve, and **(B)** loss curve of the training and validation data sets. The horizontal axis represents the number of epochs, and the vertical axis represents accuracy and loss rate, the blue line represents the validation results on the training set and the red line represents the validation results on the validation set.

**Table 3.** Predictive performance of the deep learning model.

|  | Accuracy | Precision | Recall | F1 Score |
| --- | --- | --- | --- | --- |
| HFimpEF | 0.89 | 0.98 | 0.94 | 0.96 |
| HFrEF |  | 0.94 | 0.98 | 0.96 |

## Discussion

Most of studies about deep learning-based assessment models in HF have been developed in patients with conventional HF categories, but there has been no studied in the HFimpEF patients. To our knowledge, this is the first study evaluating LVEF improvement based on deep learning algorithm, and indeed, our 2D CNN model showed good performance in HFimpEF model prediction through clinical profiles only. Our system provides an automatic interpretation that exhibits high accuracy in detecting HFimpEF. However, because of the limited amount of data available for patients with HFimpEF, our system could only provide a proof of concept for deep learning model development to aid in HFimpEF prescreening. To improve our system, further research and data collection are required. In addition, the model not only avoids premature device therapy, but also avoids invasive, expensive, and time-consuming clinical operations, reduces the burden on the patient and simplifies the process. Furthermore, this model may be able of aiding in the clinical decision making in prognosis determining by going through available patient information with less reexaminations. However, the system still has room to improve.

Similar to another study by Park et al., of 422 patients, the prevalence of HFimpEF was 28.9%^5^. We have found that the number of patients with comorbid hypertension in HFimpEF is higher than that in HFrEF. Previous study included 3,124 HF patients found that hypertension was associated with a 10% improvement in LVEF at 2.7 years follow-up^6^. In other studies, showed less comorbid hypertension in patients with HFimpEF^7^. The heart is the main target organ damaged by hypertension. Long-term higher-pressure load stimulates cardiomyocyte hypertrophy and deformation, leading to left ventricular dilatation and ventricular wall thickening, which further leads to diastolic dysfunction. A subset of patients further developing systolic dysfunction in the presence of chronic volume and pressure overload, and other patients with end-stage systolic heart failure will have more severe ventricular remodeling and less likely to have improved LVEF. We analyze if there is relevant with timely treatment of hypertension and target organ damage and conclude that target organ damage may occur if not timely treatment.

We found that the number of patients with comorbid dilated cardiomyopathy in HFimpEF is fewer than that in HFrEF. Dilated cardiomyopathy is characterized by biventricular or left ventricular enlargement and ventricular wall thinning, and myocardial structural and functional abnormalities accompanied by left ventricular systolic hypoplasia, which is a common etiology of HFrEF. A recent study by Fomin et al. found that myocardial gene mutations are the most common cause of dilated cardiomyopathy and a key path mechanism of the disease^8^. That patients with dilated cardiomyopathy are less susceptible to reverse remodeling may be associated with myocardial gene mutations and myocardial fibrosis. It was previously believed that dilated cardiomyopathy patient has a poor prognosis, with progressive reduction in LVEF and deterioration of cardiac function will ultimately lead to death. It has been found that some patients with dilated cardiomyopathy have significant improvement in LVEF after standardized drug therapy, as well as in long-term prognosis, after standardized medication therapy^9^.

Our results showed that patients with HFimpEF have lower LDL-C levels, which may be related to the inflammatory effect of LDL-C. Abnormal lipid level is either a key factor in the development of cardiovascular disease or one of the important indicators that affect the prognosis of patients with chronic heart failure. As an important indicator in blood lipids, elevated LDL-C promotes an inflammation response in the vascular lining and damages vascular endothelial cells^10^. The relationship between lipids and the prognosis of heart failure is still not understood. Other studies have suggested that elevated levels of LDL-C and TC associated to higher incidence of HF and lower survival rates.

In our study we also showed that improvement in LVEF were statistically significant to smaller baseline LVEDD and LAD. Diverse studies showed that smaller LVEDD and LAD is independent predictors of LVEF improvement in patients with HF^11,12^.

Our study found that another predictor of LVEF improvement was higher baseline prealbumin. Prealbumin offers important prognostic information in patients with HF. Jonathan et al. reported that the patients with prealbumin levels below 15mg/dl had higher morality and readmission rates, which confirms strong association with a poor prognosis in HFrEF^13^. In contrast, our results substantiate that a higher prealbumin level significantly relates to the improvement of LVEF. The underlying mechanism of this association is not well understood, higher prealbumin levels may reflect the nutritional status of a patient. There is no study have analyzed the association of prealbumin levels and LVEF improvement.

Over the past few years, an increasing number of studies concentrate on deep learning models, and most of them are based on visualizations such as ECG, echocardiogram and cardiac magnetic resonance to train models^14^. Currently, studies have been conducted domestically and worldwide to analyze diverse types of clinical data such as electrocardiograms and echocardiograms, which based on deep learning to classify and evaluate the three traditional types of heart failure and risk stratification^3,15^. However, until recently, there is no research about constructing a prediction model for HFimpEF concerning deep learning. Alkhodari et al. utilized deep learning to construct a convolutional neural network and formulated a classification model for HFrEF, HFpEF, and HFmrEF (303 cases) using clinical data. Furthermore, the convolutional neural network model derived on deep learning set up in this study predicted a 93% accuracy, the best result of any other models^3^. Based on Alkhodari et comparing the performance of the models. We have observed that the model has been optimized after adjusting the number of neurons in the convolutional layer was 64 and 128, respectively. Moreover, the present study has a larger sample size of 422 cases and faster training of the model compared to the study of Alkhodari et al.

Studies showed that the process of reverse remodeling, in patients with dilated cardiomyopathy and new-onset heart failure after initiating optimal drug therapy, may take up to 2 years, and improvement in LVEF would be longer^16,17^. Based on previously published data, the incidence of reverse remodeling ranged from 19% to 45% among patients with HFrEF^18–21^. CRT is the preferred treatment option to improve LVEF and prolongs survival. As CRT implantation is risky and expensive, the best drug therapy recommended by the guidelines is mostly used. According to the current guidelines, implantable cardioverter-defibrillator (ICD) implantation is recommended when LVEF is still less than 35% after at least 3 months of optimal drug therapy^22^. However, in patients with delayed reversal of heart failure, the timing of ICD implantation is hard to determine.

Patients with delayed improvement in cardiac function and architecture are supposed to be actively considered for guideline-guided pharmacologic therapy and avoid untimely ICD implantation. On the contrary, for patients who have highly fibrotic myocardium with little or no potential for reversal should be advised to themselves and their families as soon as possible.

In terms of study limitations, first, smaller number of non-consecutive enrollments of the patients could have potential influence of selection bias. So increasing the amount of training data set might improve the accuracy of deep learning-predicted results. Second, indicators filtered out by preprocessing may be more predicative of the model construction. Third, baseline and reviewed echocardiograms were not measured by the same physician and are inevitably biased. Fourth, as our data did not include pharmacologic profile of HF patients, it is necessary to add medicine therapy in assessment of HFimpEF.

## Nonstandard Abbreviations and Acronyms

CNN: Convolutional neural networks
DL: Deep learning
HFimpEF: Hearth failure with improved ejection fraction
HFmrEF: Heart failure with mildly reduced ejection fraction
HFpEF: Heart failure with preserved ejection fraction
HFrEF: Heart failure with reduced ejection fraction
IVSD: Interventricular septal thickness at diastole
LAD: Left atrial diameter
LVEDD: Left ventricular end diastolic diameter
LVEF: Left ventricular ejection fraction

## Data Availability

All authors have read and approved the submission of the manuscript, which has not been published and is not being considered for publication elsewhere in whole or part in any language.

https://www.clinicaltrials.gov/study/NCT06070506

